# A longitudinal study into the correlation between faecal urease activity and incidence of nappy rash in infants

**DOI:** 10.1101/2025.09.27.25336806

**Authors:** Krystal A. Le Doaré, Vicky Hunt, Robyn Deeks, Amanda Vick, A. Toby. A. Jenkins

**Author notes:** Toby Jenkins author to whom correspondence should be addressed. Krystal A. Le Doaré.

## Abstract

**Objectives:** The objective of this study was to look at whether there is a correlation between urease activity in babies’ faeces and observed incidence of nappy rash (diaper dermatitis) in a six-infant longitudinal observational study conducted at a university childcare facility over 9 months.

**Methods:** Six babies who met the inclusion criteria and who attended Westwood Nursery, University of Bath were recruited to the study with consent provided by parents following favourable ethical committee opinion from the NHS Regional Ethics Committee. Soiled nappies donated up to twice weekly were analysed for the urease activity in faecal bacteria. At the same time, Nursery staff recorded the skin condition of the nappy area of participating infants.

**Results:** A clear statistical correlation between urease expression and observed nappy rash incidence and absence of faecal urease and healthy skin was observed using Chi squared analysis (P = <0.0001).

**Conclusions:** Urease expressing bacteria were first implicated in the pathogenesis of nappy rash in the early 20^th^ century. This is the first study to show a population level correlation between nappy rash and faecal enzyme activity, which can be understood in terms of a causal chain: urease catalyses ammonia production, which directly damages skin barrier function and creates a pH environment in which secondary opportunistic micro-organisms can grow at an enhanced rate and increase skin damaging enzyme activity, therefore leading to more severe nappy rash.

**Key messages:** *What is already known:* the involvement of urease /ammonia expressing bacteria in the pathogenesis of nappy rash has been suggested for over 100 years.

*What this study adds:* This study shows a clear temporal correlation between faecal urease expression and nappy rash incidence (and vice versa) in a group of six infants followed over 8 months suggesting the key importance of urease in nappy rash pathogenesis in a relevant study population and adds to a previous causal mechanism, where urease converts urea to ammonia, which has been shown to both directly damage skin barrier function and raise skin pH.

*How this study might affect research, practice or policy:* This study raises the possibility that nappy rash can be treated more effectively by direct inhibition of urease. Several urease inhibiting strategies are currently being studied, including watercress extract and probiotic bacteria.

## 1. Introduction

Nappy rash, also known as diaper dermatitis, is a form of irritant contact dermatitis caused by prolonged exposure of skin to urine and faeces. It has been estimated that 70% of infants have at least once incidence if nappy rash during the first 24 months of their lives and although not life threatening in infants, causes significant pain and discomfort resulting in reduced sleep and stress (for babies and their parents).^1^ Nappy rash typically presents as red and inflamed skin in the nappy area with skin ulceration and obvious fungal infection seen in some cases. Nappy rash in incontinent adults is generally referred to as Incontinence Associated Dermatitis (IAD) and is arguably a more serious condition as often observed in patients with poor mobility, pressure injury and various co-morbidities and can lead to serious tissue breakdown.^2^

The pathogenesis of nappy rash is complex: human skin has not evolved to be in frequent contact with urine and faeces: doing so exposes it to a complex ecology of pathogenic bacteria. Initially these are enteric bacteria such as *Proteus mirabilis* and *Klebsiella* species which express the enzyme urease. Urease which converts urea found in urine and sweat into ammonia. Ammonia directly damages skin barrier function creating visible erythema (figure 1).^3^ Ammonia on skin also raise skin pH, damaging the natural acid mantle of the stratum corneum, creating an environment whereby faecal enzymes including lipases and proteolytic enzymes such as trypsin activities are enhanced, the growth rate pathogenic commensal bacteria including *Staphylococcus aureus* is enhanced and fungi such as *Candida albicans* are able to alter their morphology to a skin invasive form creating more severe skin damage.

**Figure 1.**
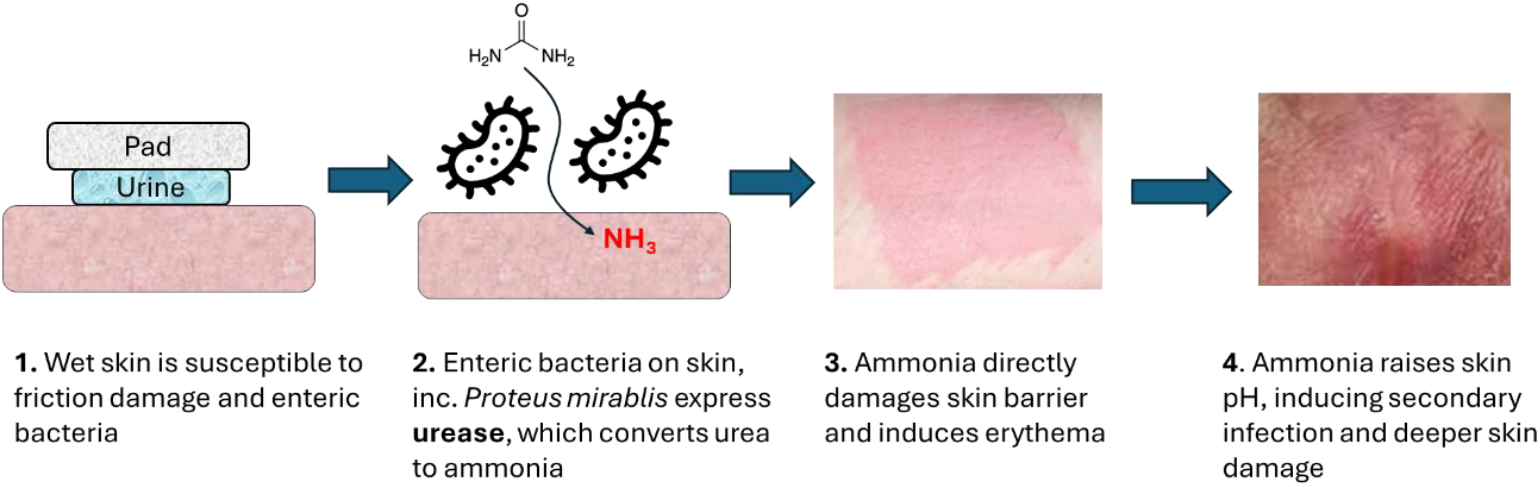
Proposed pathogenesis of nappy rash from early-stage skin redness caused by ammonia to later stage opportunistic infection and skin breakdown, showing the central role of the enzyme urease.

The association of ammonia and nappy rash was reported almost 100 years ago:^4^ Cook and Keith in 1927 reported ‘*an organism isolated in pure culture from the stools of more than 50 infants and older children involved in the splitting of urinary urea by a bacterium which infests the gluteal region and diaper from the feces’* which they named *Brevibacterium ammoniagenes*. In 1952, further bacteriological study of bacteria believed to be involved in nappy rash was undertaken by Brown, Tyson and Wilson. ^5^ They suggested that nappy rash has two phases: initially skin injury by ammonia caused by ammonia expressing bacteria followed by opportunistic infection by pathogens such as *Staphylococcus* species. However, this association between ammonia and nappy rash has been questioned, a study by Leyden et al in 1977, which attempted to measure the ammonia concentration of urine removed from the nappies of two group of infants: those who had active dermatitis and those who did not. No statistical difference in ammonia concentration was measured.^6^ A follow up paper by Berg in 1986, looking at the effect of urine and faeces on bald mice suggested that the pathogenic role of ammonia in nappy rash was via due to it increasing skin pH, and thus increasing the activity of skin damaging enzymes.^7^

A study by Owen et al in 2024, looked at the direct effect of ammonia compared with sodium hydroxide at the same pH when applied to adult human ventral forearms showed that ammonia had a directly damaging effect on skin in both causing erythema and a large reduction in stratum corneum barrier function, as measured by impedance spectroscopy, whereas the sodium hydroxide had no effect.^3^ This finding raised important questions about the role of urease and ammonia in nappy rash. A paper by Kohta et al published in 2023 reported a strong association between the urease activity of genital skin swabs of incontinent stroke patients and incidence of incontinence associated rash in those patients.^8^ This was an interesting observation as the effect of urease on skin would be to express produce ammonia *in-situ*. Which led to the question being addressed in this study: Is there a temporal correlation between urease expression from faecal bacteria and nappy rash/IAD incidence?

In this paper we report a small longitudinal study of infants (age 6-18 months) at the University of Bath childcare facility, Westwood Nursery. Our start point was to ask that given that babies’ skin is constantly exposed to urine and faeces, why don’t babies suffer from nappy rash all the time? The hypothesis being tested was that it is the urease expression levels of faecal bacteria to which babies are exposed to which is associated with incidences (or absence) of nappy rash. The rationale is that if we see an association, it further supports the urease / ammonia hypothesis suggested by Owen et al and the observations of adult IAD made by Kohta et al.^4,9^

## 2. Methods

Study protocol and participant information sheet are included in the Electronic Supporting Information.

### 2.1 Study design

#### Objectives

This longitudinal cohort observational study has the following primary objectives: (1) To look at correlation with the faecal flora, and specifically urease-expressing bacteria, of infants in the study and incidences of nappy rash over the 8-month study period. (2) To look at recorded nappy rash incidence in infants and relative urease activity in their faeces over the 8-month period.

#### Participant selection

12 infants were eligible for inclusion, 7 were initially recruited, although due to insufficient soiled nappies during the nursery day, one infant was excluded, six were followed through the course of the study, up to 8 months.

#### Consent and ethics

Parents of eligible babies were contacted by Nursery staff and given a detailed patient information sheet (see ESI) before being asked to formally consent to participation of their child in the study. No payment of other incentive was given to participating parents. Participating infants were given anonymous IDs: the scientific team had no knowledge of the true identities of babies or their parents. Ethical permissions were given by the NHS Regional Ethics committee: PR committee REC reference 23/NW/0390; IRAS project ID: 332482.

#### Inclusion and Exclusion criteria

Inclusion Criteria: infants aged 6-18 months at study start with parents willing to consent; infants who wear nappies (diapers) 24h / day; infants who wear disposable nappies. Exclusion criteria: Any infant whose parents did not consent; any infant who, at Nursery, wears reusable nappies; any infant whose parent’s English language was not sufficient to properly understand the study.

#### Study methodology

Infants taking part in the trial had their soiled (faecal) nappies placed in a ziplock bag containing the infant’s anonymous participant ID as part of their normal care routine. Nursery staff would record the skin condition of the skin in the nappy area (see ESI figure 1). The form was added to the ziplock bag and soiled nappies would be collected by scientific staff at 17:00 on study days (Tuesdays and Thursdays). Faecal matter (ca. 2 grams) was placed into Eppendorf tubes in the laboratory and flash frozen in liquid nitrogen before being stored at -20 °C until required. Eppendorf tubes were labelled with the infant ID number and collection date to allow for traceability of samples. Figure 2 illustrates the study methodology. Participant Information Sheet and Study Protocol are supplied in the ESI

**Figure 2:**
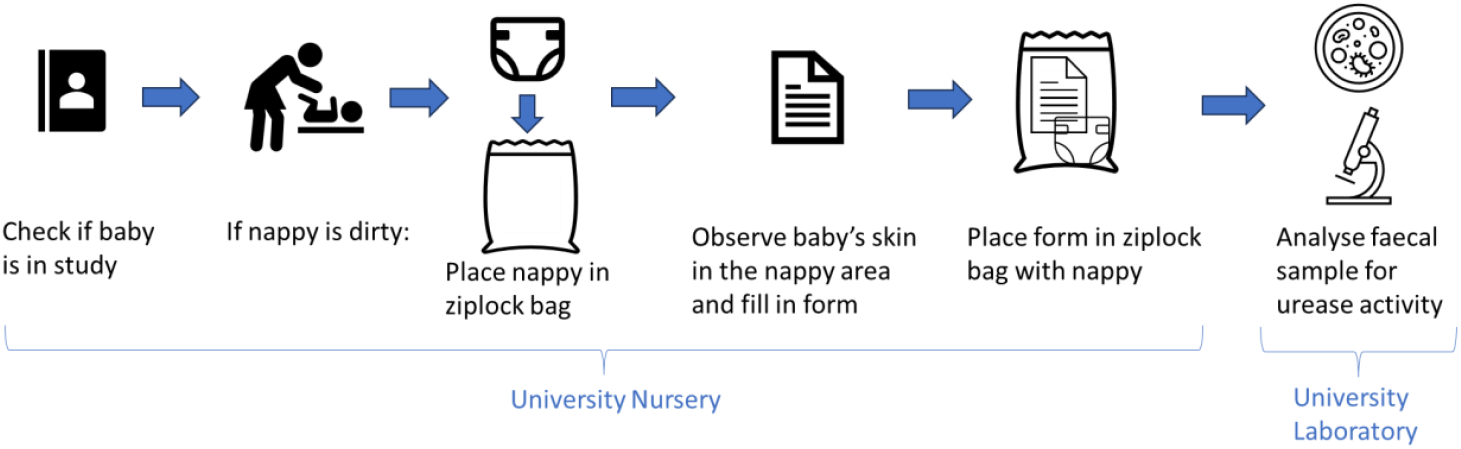
Schematic of study methodology, sample collection and analysis of faecal samples

### 2.2 Sample processing

Urease activity was quantified by thawing the Eppendorf tube, weighing out 250 mg of faecal material and resuspending in phosphate buffered saline (1 mL) and vortexed for 10 seconds. Samples were then sonicated in an Ultrawave sonicator for 1 minute before vortexing for 10 seconds to ensure sample homogenisation. Homogenised samples (50 µL) were then spread on urea agar using a method adapted from Kohta et al.,^9^ and incubated at 37 °C for 48 hours. The urea agar contains the pH indicator, phenol red, which turns to from yellow to a bright pink colour at pH >7.4 (figure 3), qualitatively indicating urease activity, as well as nutrient for bacterial growth and urea substrate for bacterial urease. Each faecal sample was cultured on three separate culture plates. A total area of pink colour change of minimum 80% of plate area was taken as the lower threshold of positive urease activity.

**Figure 3a:**
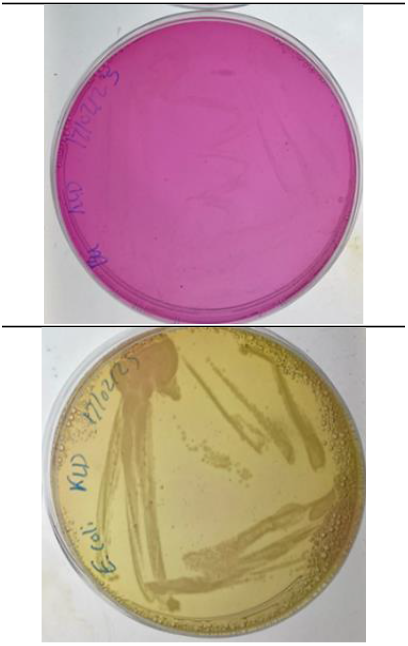
Validation of phenol red urea agar showing urease-expressing *Proteus mirabilis* B4 (top); non-urease expressing *E. coli* (bottom)

**Figure 3b:**
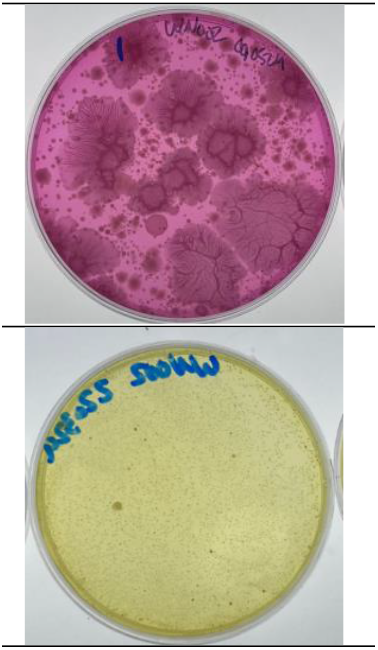
Phenol red urea agar response to infant faecal sample containing urease expressing bacteria (top) and non-urease expressing bacteria (bottom)

### 2.3 Patient and public involvement

Parents of young children were involved in preliminary studies used to validate the study protocol. The research question and outcome measures were developed in discussion with both Nursery staff and parents of young children at the Nursery. Study design involve discussion with Nursery staff to ensure it could be undertaken with minimal disruption to the working of the baby room in the Nursery. Recruitment was made via posters within the Nursery and word of mouth discussion with parents arriving to collect their children. Posters of the study findings were made and displayed in the Nursery on the advice of staff.

## 3. Results and Discussion

### Temporal association of nappy rash and faecal urease activity

Nappy rash incidence and faecal urease expression plotted as a function of study time (March – October2025) was plotted for all six participants in the study and is shown below in figure 4. Most babies showed a clustering of urease activity with nappy rash observation, and not the random distribution one might expect if the two events were not correlated. Five of the six babies (WN002; WN003; WN004; WN005; WN007) all show a clear apparent temporal association between faecal urease activity and observed nappy rash. Baby WN006 showed the least apparent correlation with 7 measured episodes of high faecal urease activity but experienced only one reported nappy rash episode. In most cases, elevated urease expression was observed in the week preceding, or the same time as the observed nappy rash episode.

**Figure 4:**
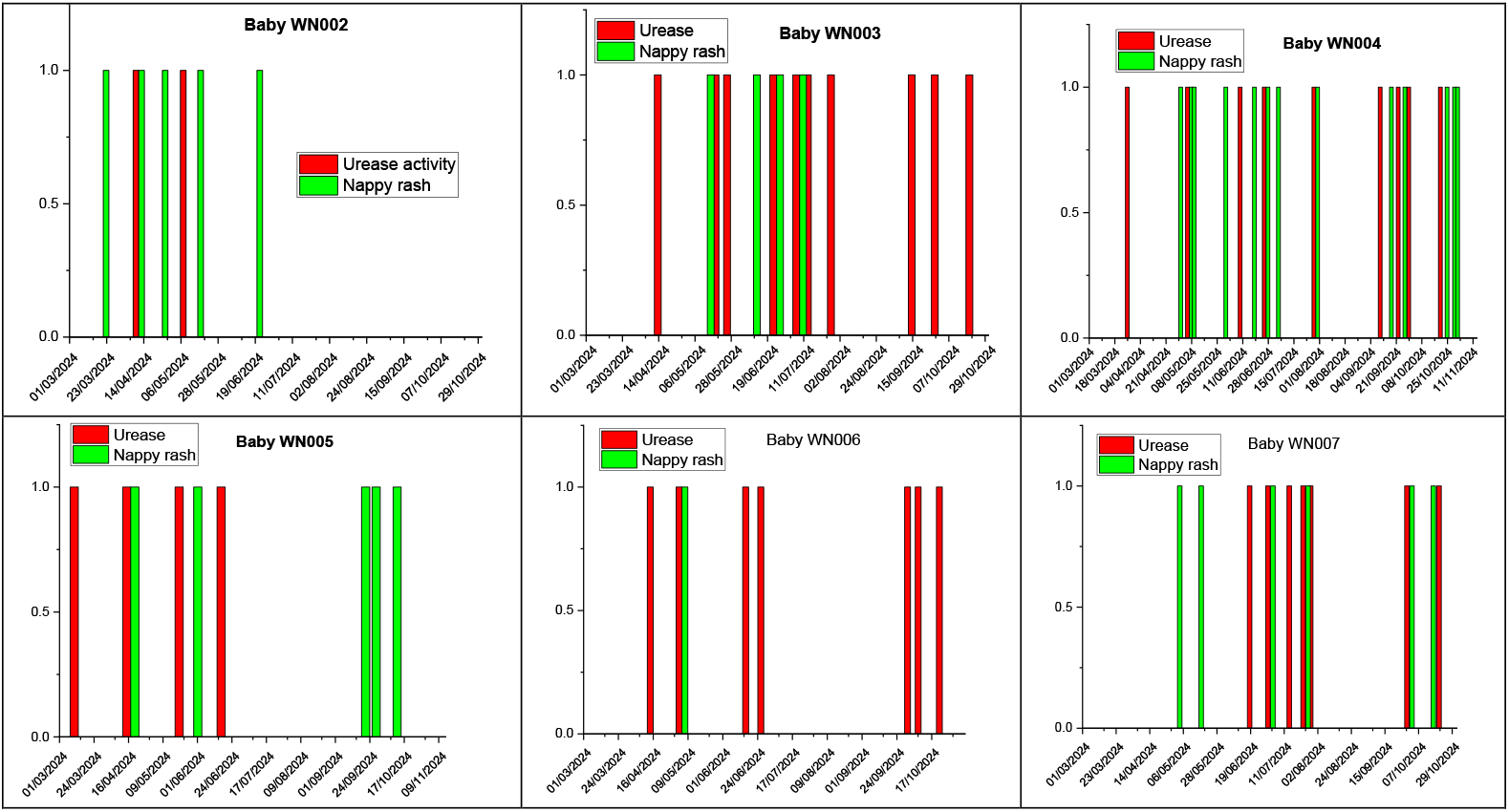
Temporal correlation between reported diaper rash (nappy rash) and urease expressing faecal bacteria for the 6 babies in the study

### Statistical analysis

Correlation was deemed to be positive if faecal urease and nappy rash if nappy rash was observed up to 7 days preceding nappy rash incidence. The reports of nappy rash and urease activity (yes/no) are both non-parametric, so correlation was quantified using Chi-squared analysis using GraphPad. The results grid is shown in table 1, positive correlation in green, negative in red:

**Table 1:** Results from all measurements (individually reported nappy rash and faecal urease of same baby at same time) of the six babies in the study looking at correlation between reported nappy rash and urease positive faecal bacteria and vice versa.

|  | Observed nappy rash | Normal skin |
| --- | --- | --- |
| Urease positive faecal bacteria | 29 | 18 |
| Urease negative faecal bacteria | 12 | 70 |

Analysis using GraphPad of the Chi-square without Yates correction gave a Chi squared equal to 30.5 with 1 degree of freedom and a two-tailed P value of <0.0001. The association between faecal urease activity and presence of nappy rash (and vice versa) was found to be extremely statistically significant.

### Discussion

The study results suggest that there is temporal relationship between faecal urease and observed nappy rash. This can be mechanistically understood in terms of direct ammonia damage to skin from the urease expressing bacteria and supports with both Cooke and Keith’s hypothesis from 1927 and more recent work showing how ammonia on skin directly damages the stratum corneum barrier function.^4,3^ Ammonia is a weak base, which increases skin pH, which also promotes the growth of pathogenic bacteria and enhances the activity of skin degrading enzymes, so has a dual role of nappy rash pathogenesis. Urease expressing bacteria in faeces was detected only intermittently, indeed the correlation between children’s healthy skin and the *absence* of measurable urease in their faeces was also significant. The putative role of urease expressing bacteria and ammonia damage to skin is surprisingly absent from much of the more recent literature. In a review from 2022 by Petek et al of emerging links between infant microbiome and skin immunology in nappy rash no mention of urease is made, and ammonia is only discussed in the context of how it raises skin pH and enhances the activity of faecal enzymes.^9^ Zheng reported that certain Phyla such as *Proteobacteria* and *Enterococcus* which are both known to contain some species which are urease-expressing, and have a significant increase in activity in infants with nappy rash than when compared to normal infant skin samples which correlates with our proposed mechanism.^10, 4^

Urease is an enzyme expressed by several enteric bacterial species, including *Proteus mirabilis*. It seems reasonable to assume that many of the skin dwelling bacteria found in the nappy area of infants have an enteric origin, which creates the possibility of a future approach for preventing nappy rash: inhibition of bacterial urease and/or inhibition of urease expressing bacteria. This might be achieved in various ways: alteration of the infant gut microbiome by dietary modification – for example by supplementation of infant formula with probiotics, direct topical application of urease inhibitors either as a cream or by modification of the skin facing top sheet of nappies. Current nappy rash topical creams work primarily via an emollient and skin barrier effect, and in some cases a weak antimicrobial effect from zinc oxide.

Inhibiting urease may have multiple effects: 1. by reducing ammonia expression and a skin ammonia concentration, skin stratum corneum integrity can be better maintained and the skin should be less susceptible to damage via hyperhydration and friction; 2. Reducing ammonia concentration will ensure skin pH can be maintained at healthy levels around pH 5.5 and crucially below pH 7, at which pH many skin damaging faecal enzymes become much more active; 3. High pH (> pH 8) is known to change the morphology of commensal yeasts such as *Candida albicans* changing the shape from a relatively harmless spherical morphology to a hyphae growing morphology which is known to be associated with the ability of such yeasts to penetrate into the skin epithelium.^11^

## 4. Conclusions

The study described in this communication does have some limitations: principally the small sample size and the subjective diagnosis of nappy rash by Nursery staff. This was not a clinical environment, and it is likely that there would be some variability in diagnosis based on staff experience and pre-conceived notions by the staff of the presentation of nappy rash. Ideally, clear photographs of the babies’ nappy area would have been taken, with anonymous scoring of skin condition by 3^rd^ parties, but this would have made receiving parental consent more difficult. The differential diagnosis for nappy rash from other dermatological conditions such as atopic dermatitis and psoriasis is discussed by Fölster-Holst.^12^ It is suspected that there may have been some over-reporting of nappy rash incidence by Nursery staff (‘*if you look for a condition, you find it’*), but this is very hard to control for without a more rigorous diagnostic criteria using photographs of infant skin in the nappy area assessed by 3^rd^ parties.^13^

The further limitation is the on/off nature of the urease detection assay. The urea agar plate assay was easy to use and gave clear results, but data was inherently qualitative. In principle a more quantitative assay could have been used, but this would have made analysis considerably more complex: a pilot study using this approach showed very high variability in results, probably due to the difficulty in standardising faecal samples.

Despite these limitations, this study is the first (to our knowledge) which clearly shows a clear faecal urease – nappy rash correlation and the absence of faecal urease correlated with absence of observed nappy rash. This likely suggests that urease does indeed have a role to play in the onset of nappy rash. In future, analysis of exactly which faecal bacteria are causing these observations may assist in the understanding of the role of urease in nappy rash is currently underway involving 16S sequencing of faecal bacteria and will be reported in due course.

## Acknowledgements

We would like to thank all Westwood Nursery staff involved in the study who packaged up soiled nappies and recorded babies’ skin condition.

## Informed consent

All parents gave full informed consent for inclusion of their children in the study. Participating parents were consented following review of participant information sheet.

## Ethics

Ethics approval statement: Ethical permissions were given by the NHS Regional Ethics committee: PR committee REC reference 23/NW/0390; IRAS project ID: 332482.

## Data availability

Data is provided in the electronic supporting information and from the corresponding author.

## Author Contributions

- Krystal Le Doare: Sample laboratory analysis, data analysis, drafting of manuscript.
- Vicky Hunt: Study design and data analysis.
- Robyn Deeks: Data analyisis
- Amanda Vick: Recruitment of infants and sample and data collection.
- Toby Jenkins: Study design, study conceptualisation, data analysis, manuscript drafting.

## Graphical abstract

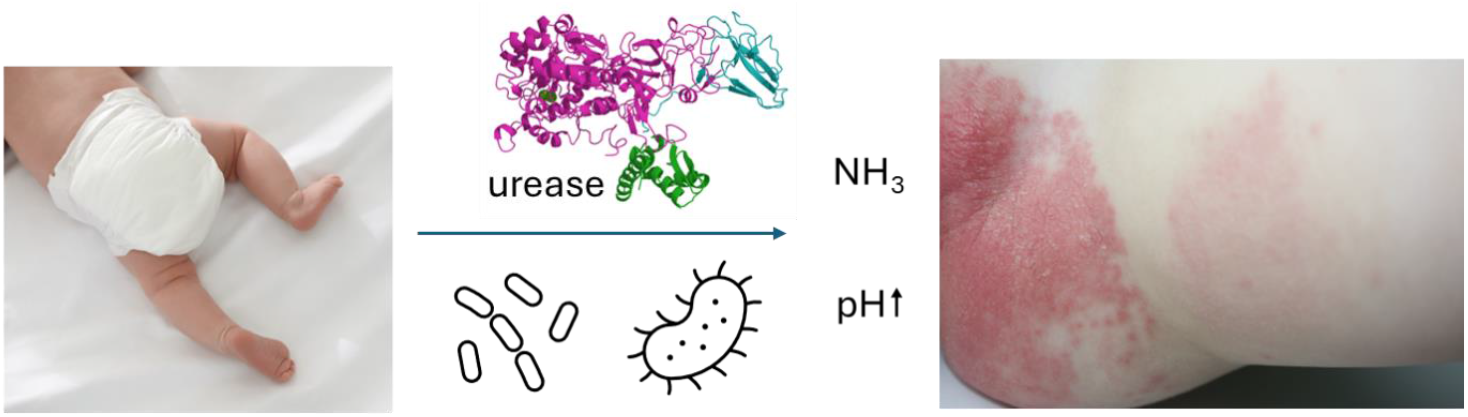

