## Supporting Information for "A longitudinal study into the correlation between faecal urease activity and incidence of nappy rash in infants"

1. Department of Chemistry, University of Bath, Bath, BA2 7AY, United Kingdom
2. Department of Life Sciences, University of Bath, BA2 7AY, United Kingdom
3. Westwood Nursery, University of Bath, Bath, BA2 7AY, United Kingdom
3.

Krystal A. Le Doaré:
Vicky Hunt:
Amanda Vick:

#### 1. Participant Information Sheet

##### Introduction

We would like to invite you to take part in our research study. Before you decide, it is important that you understand why the research is being done and what it would involve for you and your child. Please take time to read this information, and discuss it with others if you wish. If there is anything that is not clear, or if you would like more information, please ask us.

##### What is the purpose of the study?

This study seeks to better understand the relationship between the bacteria in babies faeces and incidences of nappy rash, over a 10 month period.

##### Why have I been invited?

You have been invited as your child is in the baby room at Westwood Nursery and is 6-18 months and wear nappies full time. We are looking to enrol around 12 babies in this study over a period of 10 months.

##### Do I have to take part?

The answer is 'No': Taking part is entirely voluntary. You can withdraw your baby at anytime if you change your mind without giving a reason; Withdrawal will not affect the care your child receives in any way.

###### What will happen to me if I decide to take part

At the start of study your baby's age, history of skin conditions i.e. eczema, and feeding regime (formula, breast milk, solid food) will be recorded and the baby given an anonymous ID number. The study will last for up to 10 months. On Mondays and Thursdays (both days if your child is in the nursery on both day, otherwise on one of these days):

When your child has a soiled (dirty) nappy, the nappy will be placed in a ziplock bag with their anonymous ID affixed to the bag.

A short questionnaire on the baby's skin health in the nappy area will be filled in with their anonymous ID attached and placed in the bag, with the nappy. The nappy and form in ziplock bag will be collected on Mondays and Thursdays at 17.00-17.30 by the scientific team. **No** photographs of the baby's skin will be taken.

The schematic below illustrates the process, as far as it involves your baby directly:

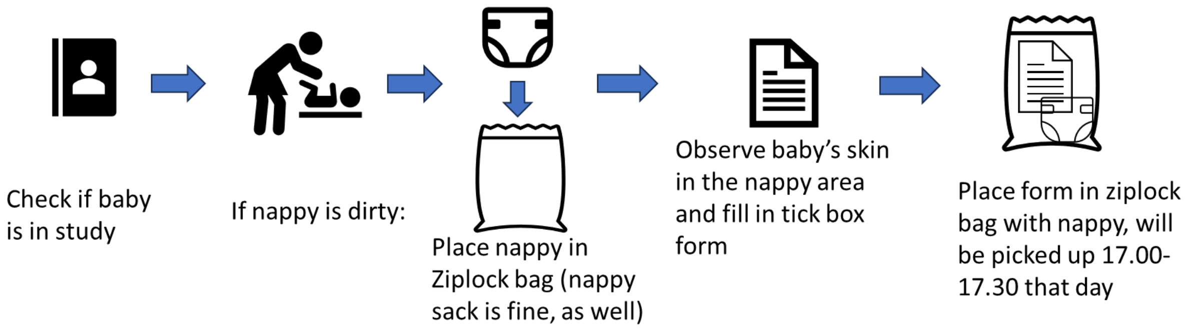


**Are there any possible disadvantages or risks from taking part?**

We cannot see any disadvantage for any child/parent to participate: they will not receive any extra or less care; no human DNA will be analysed or stored; your child will be anonymous; your child will not suffer any pain or discomfort from taking part.

**What are the possible benefits of taking part?**

There is no direct benefit for your child, but it could help develop better targeted treatment for nappy rash which could benefit children in the future.

**Will my General Practitioner/family doctor (GP) be informed of my child's participation?**

No.

**Will my taking part in the study be kept confidential?**

All babies will be given an anonymous identifier with only baby room staff having knowledge of which ID correlates to which baby. This information will never leave the baby room at Westwood Nursery, and scientific staff will have no idea of babies / parents real identities. At end of study, the identifier key will be destroyed. It will not be possible to withdraw data relating to your child before the date of withdrawal.

**Will I be reimbursed for taking part?** No reimbursement is possible.

**What will happen to the sample and questionnaire?**

Over the 10 months of the study, faeces in the nappy will be analysed in three ways: 1. We will analyse the faeces to identify which bacteria are present; 2. We will measure the activity of the enzyme urease (which converts urea to ammonia); 3. We will measure the pH (acidity) of both the faeces and urine in the nappy. We will look at the correlation between changes in faecal bacteria, urease activity and pH with nappy rash incidence. Results will be correlated with the baby skin health questionnaire, which will have been filled in by nursery staff. Samples will be disposed of after analysis.

**What will happen to the results of this study?** Results will be disseminated via peer-reviewed scientific publication and talks at conferences such as The European Society for Paediatric Infectious Disease annual meeting. None of the participants will be identifiable, all data will be anonymised. Results will also form part of the doctoral thesis of the PhD student, Krystal Le Doare.

**What if there is a problem?** The Nursery team will consult with parents in the usual way in case of any concern regarding your child's health. If you have concerns contact the Chief investigator:

**Data security**

**How will we use information about you?**

We will need to use information from your child for this research project.

This information will include your child's:

· Name and date of birth

· Weekly / bi-weekly assessment of your child's skin

· Results from the scientific investigation which relate to your child's skin health including faecal bacteria and urease enzyme activity.

Information about the name and date of birth of your child will be held securely at Westwood Nursery – scientific staff will **not** have access to this information. Information on your child's skin health and scientific data will be held securely in the Department of Chemistry, University of Bath.

People will use this information to do the research or to check your records to make sure that the research is being done properly. People who do not need to know who you are will not be able to see your child's name or contact details. Your data will have a code number instead.

**We will keep all information about your child safe and secure.**

**What are your choices about how your information is used?**

You can stop being part of the study at any time, without giving a reason, but we will keep information that we already have.

**Where can you find out more about how your information is used?**

You can find out more about how we use your information

at www.hra.nhs.uk/information-about-patients/

our leaflet available from www.hra.nhs.uk/patientdataandresearch

by ringing us on 01225 386118

**Who is organising and funding the study?**

The study is funded by The Annette charitable trust, The Rosetree foundation and the University of Bath. The study is organised and run by the chief investigator, Professor Toby Jenkins.

Who is involved in this study:

**Lead Investigator at University of Bath:** Krystal Le Doare:

**Principal investigator at Westwood Nursery:** Mrs Amanda Vick:

**Chief investigator at University of Bath:** Professor Toby Jenkins:

**Sponsorship of the study:** The University of Bath is sponsoring the study

Reviewed and approved the study : PR committee REC reference 23/NW/0390 ; IRAS project ID: 332482

#### 2. Baby skin health questionnaire

**Unique ID of baby** (do not write name):
**Date:**

**Baby's diet at this time (if known):**
Solid food and breast milk ☐;
Solid food and formula milk ☐;
Breast milk only ☐;
Formula milk only ☐

**Signs of baby teething (pain, redness etc)?**

Yes ☐ /No ☐ / Maybe ☐

Condition of skin in nappy area:
Normal ☐;
Red in places ☐;
Extensive redness ☐;
Redness and skin breakdown ☐;
Evidence of yeast infection ☐;

**Baby has nappy rash?** Yes ☐ /No ☐ / unsure ☐
**Any other observations?**

### Gut Microbiome & Skin Health Study

**Version Number: 4.5**

**Dated: 6 December 2023**

**REC Number:** 23/PR/1497

**IRAS Number**: 332482

**Sponsor Name & Address:** University of Bath

**Funder:** The Annette Trust

**Planned Study Period: 15 January 2024 – 20 December 2024**

**Protocol authorised by:** Professor Toby Jenkins, Chief Investigator

**Date:** **6 December 2023**

### CI and Research Team Contact Details

**Lead investigator**
Ms. Krystal Le Doare
Department of Chemistry
University of Bath
Bath BA2 7AY


**Chief Investigator:**
Prof. Andrew Tobias Aveling 'Toby' Jenkins
Professor of Biophysical Chemistry
Department of Chemistry
University of Bath
Bath BA2 7AY


**Baby room leader (site lead) at Westwood Nursery:** Mrs Amanda Vick, Baby room lead. Westwood Nursery Manager, University of Bath, BA2 7AY


**Co-Investigators:**
Dr Kyle Stewart, Corner Place Surgery, 46A Dartmouth Road, Paignton, TQ4 5AH
Dr Vicky Hunt, Department of Life Science, University of Bath, BA2 7AY
Dr June Mercer-Chalmers, Research & Innovation Services, University of Bath, BA2 7AY

**Details of Sponsor:**
The University of Bath is the research sponsor for this study.

**Funder:** This study is being funded by the Annett Charitable Trust

This protocol describes the Gut Microbiome and Skin Health (GuMeSH) study and provides information about procedures for entering participants. Every care was taken in its drafting, but corrections or amendments may be necessary. These will be circulated to investigators in the trial. Problems relating to this study should be referred, in the first instance, to the Chief Investigator.

This study will adhere to the principles outlined in the UK Policy Framework for Health and Social Care Research. It will be conducted in compliance with the protocol, the Data Protection Act and other regulatory requirements as appropriate.

**Abbreviations**

| | |
|---|---|
| UoB | University of Bath |
| PIS | Participant Information Sheet |
| AE | Adverse Event |
| AR | Adverse Reaction |
| SAE | Serious Adverse Event |
| SOP | Standard Operating Procedure |

**Keywords:** dermatitis, nappy rash, faecal microbiome

#### Table of Contents


| | | | |
|---|---|---|---|
| **Chief Investigator (CI) and research team contact details** | | | 6 |
| **1** | **Introduction** | | 8 |
| **2** | **Study aims and objectives** | | 9 |
| | **2.1** | Study aims | 9 |
| | **2.2** | Study objectives | 9 |
| | **2.3** | Participant selection | 10 |
| **3** | **Study design** | | 11 |
| | **3.1** | Recruitment and study pathway | 11 |
| | **3.2** | Study design | 11 |
| | **3.3** | Sample processing | 12 |
| | **3.4** | Sample disposal | 12 |
| | **3.5** | Nursery follow up | 12 |
| | **3.6** | Data security | 12 |
| **4** | **Participant entry** | | 13 |
| | **4.1** | Inclusion criteria | 13 |
| | **4.2** | Exclusion criteria | 13 |
| | **4.3** | Data collection and analysis | 13 |
| | **4.4** | Sample size | 13 |
| | **4.5** | Results dissemination | 13 |
| **5** | **Reporting** | | 13 |
| | **5.1** | Safety reporting, Adverse Events (AE) or Adverse Reactions (AR) (AE/AR) | 13 |
| | **5.2** | (Serious) Breaches | 13 |
| | **5.3** | Progress reporting and end of study reporting | 13 |
| | **5.4** | Final report and dissemination to parents and Nursery staff | 14 |
| | **5.5** | Peer reviewed scientific publication | 14 |
| **6** | **Regulatory reviews** | | 14 |
| | **6.1** | Authorisations and Research Governance Statement | 14 |
| | **6.2** | Consent | 14 |
| | **6.3** | Confidentiality | 14 |
| | **6.4** | Indemnity | 14 |
| | **6.5** | Sponsor | 15 |
| | **6.6** | Funding | 15 |
| | **6.7** | Monitoring and audit | 15 |
| | **6.8** | Study management | 15 |
| **7** | **Publication policy: Peer-reviewed scientific journals, internal reports and conference presentations.** | | 15 |
| **8** | **References** | | 15 |
| **9** | **Appendices** | | 16 |

### 1.0 Introduction

Urease is a nickel-based metalloenzyme expressed by various bacteria, including gut dwelling *Proteus mirablis*, which converts urea to ammonia. The role of urease-expressing *P. mirabilis* in the aetiology of ‘ammonia dermatitis’ / nappy rash was first reported almost 100 years ago.[1] Urease expressed by *P. mirabilis* is associated with the pathogenesis of invasive incontinence-associated dermatitis (IAD or “nappy rash”). Urease converts urea to ammonia, which is directly skin irritating, reducing the stratum corneum barrier function and activating proteolytic enzymes whilst raising pH, promoting bacterial and fungal growth (figure 1).[2] On skin, high pH promotes the formation of hyphae (the long, thread-like filaments or tubes in fungi) in *C. albicans*, which can cause tissue damage by invading mucosal epithelial cells, leading to blood infection.[3]

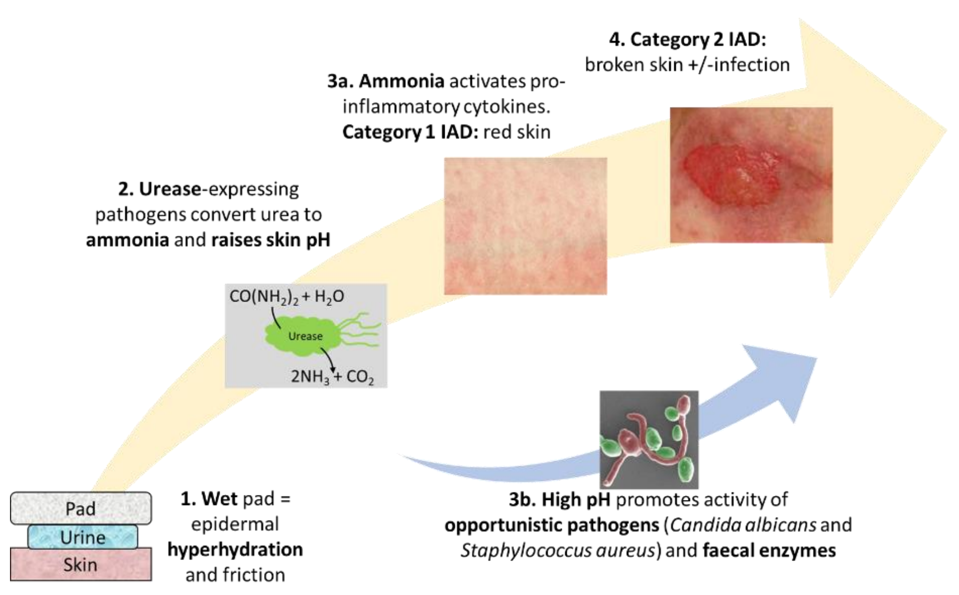


**Figure 1:** Multiple micro-organisms and enzymes are involved in the complex pathogenesis of IAD/nappy rash, but the onset of IAD is associated with urease, which converts urea to ammonia.

Currently, nappy rash is treated by applying a barrier cream to the skin, creating a physical barrier between the urine / faeces in the nappy and the skin, whilst providing a mild antiseptic function. Frequent nappy changing along with keeping the skin clean, is also effective. However, there are no interventions which specifically target the primary virulence factor, urease.

#### Work to date by the UoB laboratory

##### *Skin barrier damage by Proteus mirablis on in-vivo human skin nappy rash model*

The team at UoB are developing both ex-vivo and in-vivo (human forearm) nappy rash models, whereby skin is soaked in artificial urine, then inoculated with *Proteus mirablis* and changes in skin barrier function measured using impedance spectroscopy.[2] Impedance spectroscopy is very sensitive to changes in the stratum corneum barrier function. Figure 2 shows data from human forearm skin resistance following inoculation with artificial urine, artificial urine plus *Proteus mirablis* (wild type) and a urease negative mutant of *P. mirablis*. It can be clearly seen that, after 4 hours, the urease expressing *P. mirablis* raised skin pH to 8 and reduces stratum corneum resistance by 80%.

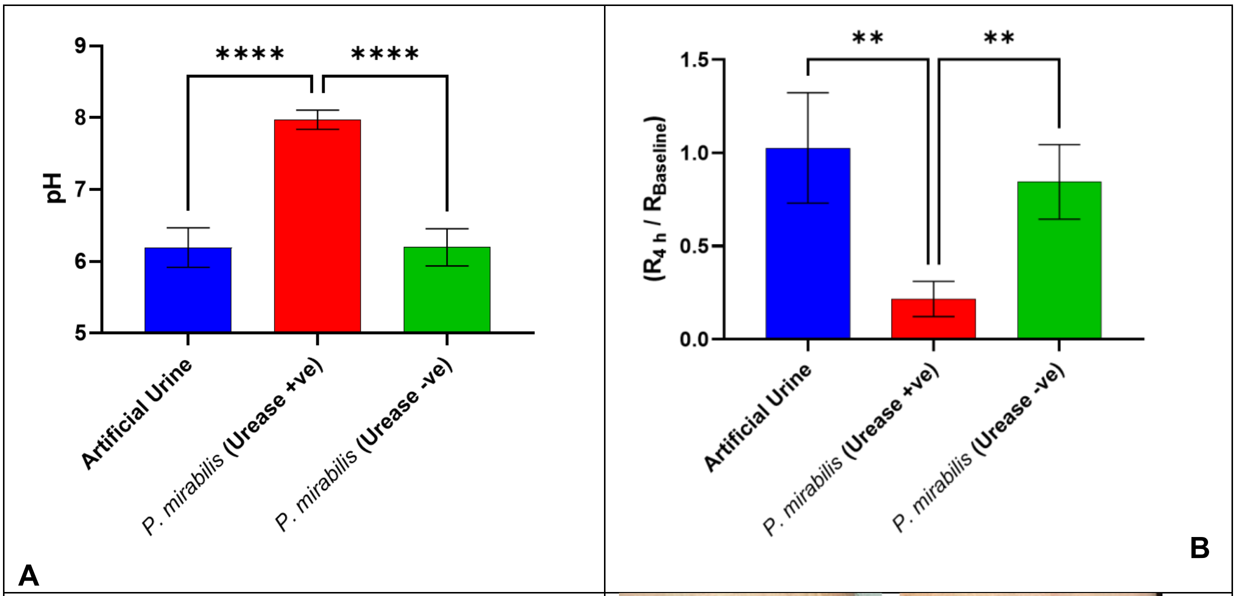


**Figure 2, A.** In-vivo skin pH following 4 h inoculation with artificial urine, urease positive and negative *P. mirablis.*

**B.** Stratum corneum resistance at 4 hours: no significant change for artificial urine and urease negative *P. mirablis*.

**C.** Photograph of erythema on human forearm following urease positive *P. mirablis* inoculation.

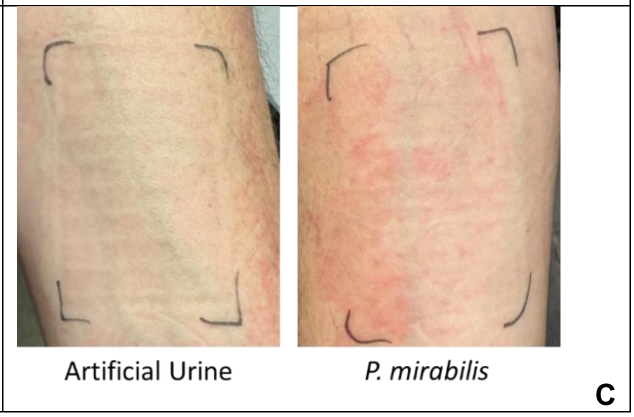


### Study aims and objectives

#### 2.1 Study Aims

The key research question is: why don't babies suffer from nappy rash all the time, given that their skin is exposed to urine and faeces daily? Anecdotally, episodes of nappy rash get worse when babies are teething—but is this true and, if so, why should this be?

The team intends to test the hypothesis that incidences of nappy rash in babies correlate with changes in their faecal microbiome and with elevated levels of urease / urease expressing bacteria in their faeces.

##### 2.2. Objectives

**Primary objectives:** (1). To look at correlation with the faecal flora, and specifically urease-expressing bacteria, of babies in the study and incidences of nappy rash over the 10-month study period; (2). To look at recorded nappy rash incidence in babies and relative urease activity in their faeces over the 10-month period; (3). To look at changes in levels of other pathogenic microorganisms, including *Staphylococcus aureus* and *Candida albicans,* and nappy rash incidence / severity.

**Secondary objectives:** To look for any correlation with babies' diet and incidence of teething with nappy rash incidence / severity; to measure pH in the nappy and correlation with skin health.

**Study Design:** a longitudinal cohort observational study

***Study group***: babies in the baby room at Westwood nursery, Claverton Down Campus, University of Bath.

***Study duration for each child:*** 10 months; up to 2 soiled nappies to be donated per week.

***Criteria for discontinuing:*** parents withdraw baby from study; child leaves the nursery.

##### 2.3 Participant selection

***Expected number of eligible participants available per year and proportion of these expected to agree to the study:*** 12 babies.

***Provision of participant information sheet***: Parents will be given the Participant Information Sheet (PIS) at start of study.

***Gaining participants' consent:*** Baby room lead and Principal Investigator (Mrs Amanda Vick or deputies) will consent parents following provision of the PIS and a verbal description of study and inclusion / exclusion criteria applied. The Lead Investigator (LI) Krystal Le Doare and (CI) Chief Investigator (Toby Jenkins) will run an on-line (Teams) meeting for interested parents following initial contact with parents by Amanda Vick. This will give scientific background and answer any questions with respect to ethical issues, data use etc.

***Detail of enrolment procedure:*** Parents will be approached by nursery (baby room) staff at beginning, or end, of nursery day - when dropping off for collecting their children. Following the Teams meeting run by the LI and CI, parents will be invited to give informed consent by the baby room lead, Mrs Vick, or her deputies.

***Subject compliance:*** Provided babies are consented, fit to inclusion criteria and are present in the nursery on the relevant days, they will participate until they either leave the baby room, leave the nursery or parents choose to withdraw, or the date is after 24 December 2024.

***Withdrawal:*** Parents may withdraw their child at any time, either by verbal instruction or email. Unless specifically requested, samples from babies will be processed up to the withdrawal date.

***Sample size calculation:*** This is a pilot study, sample size based on size of baby room population and resources available.

***Data collection:*** The only data collected will be the form depicted in appendix 1 and the microbial data of their faeces: bacterial / fungal species and abundance, urease gene expression levels and pH. Data will be collected up to twice weekly, until study end.

### Study design

#### 3.1 Recruitment

**Subject recruitment**
***Method of recruitment:*** Poster in nursery and flyers; word of mouth from baby room staff.

***Payment of participants:*** No payment or expenses

***Details of procedures, tests, and screenings carried out to assess study suitability:*** Nursery staff will screen babies with respect to inclusion / exclusion criteria. Each participating baby will have a folder with their name and their anonymous ID number written on it, kept in nappy changing area. Scientific staff will not have access to this folder.

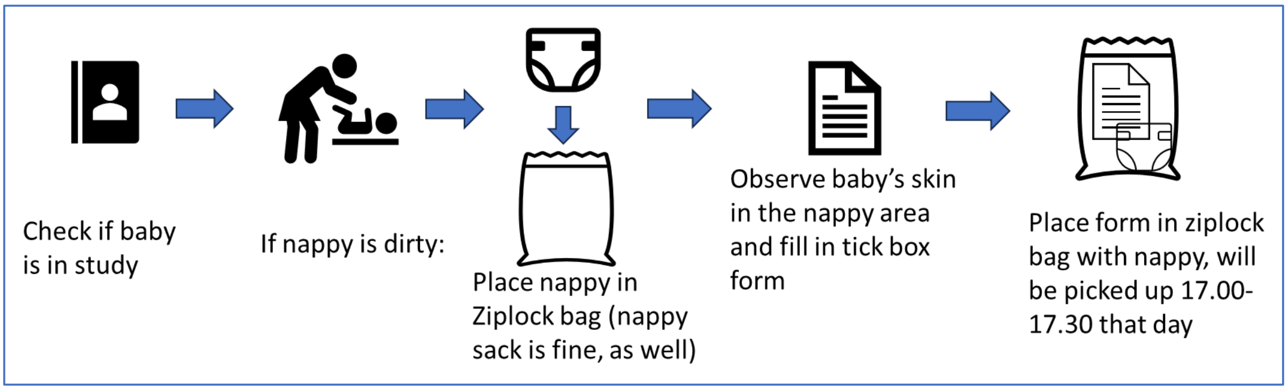


**Figure 3:** schematic of procedure from perspective of baby / nursery staff

#### 3.2 Study design

***At start of study:*** Nursery staff will record baby's age, history of skin conditions i.e., eczema, and feeding regime (formula, breast milk, solid food) will be recorded and the baby given an anonymous ID number (figure 3).

**Note:** Not all babies will attend nursery on Monday and Thursdays. Babies only present on Monday or Thursday are still eligible, but will only donate one nappy per week. Due to holidays etc, there will be weeks where some data on participating babies is not collected.

*On Mondays and Thursdays:*

- On changing dirty nappies (as part of routine care), put nappy into Ziplock bag with anonymous ID on bag for that baby.
- Fill in tick box form for that baby, put form in Ziplock bag with nappy (Appendix 1).
- Put nappy/form in Ziplock bag in special bin: will be collected 17.00-17.30 by the scientific team.
- Nappies will be brought to the laboratory and stored in a dedicated fridge prior to (next day) removal of around 5 g of faecal material and testing of pH of soiled part of nappy with flatbed pH probe. The baby's study ID number and date of collection will be recorded on all samples, to allow traceability of samples.

#### 3.3 Sample processing

All samples will be taken using a standard method: Collected faeces (from nappy) will have DNA extracted using a standard kit and either processed immediately or frozen at -20°C until processing. Only bacterial and fungal DNA will be analysed. Human DNA will not be studied or analysed.

Faeces will be studied in three ways:

1. Urease activity (measured in International Units (IU) / mg of faecal material) will be quantified using the Berthelot assay. Our hypothesis is that nappy rash incidence will correlate with elevated faecal urease activity, so we will look at bi-weekly urease activity measurement with the reported skin health of babies in the study.
2. Bacterial species: faeces contain many bacterial species, including *Proteus mirablis*, which is known to have high urease expression levels. As with urease, will study how change in the bacterial species flora correlates with reported skin health of the babies.
3. pH of the skin contact area I the nappy will be measured using a flat-bed pH probe, again high pH (>7.5) is expected to correlate with nappy rash incidence.

Bacterial and fungal DNA will be gene sequenced (16S/18S rRNA) and analysed by MALDI-ToF to determine which primary bacterial species are present. Babies will be flowed longitudinally, changes in baby's diet and incidences of teething also being recorded to give a more wholistic picture of how these factors might correlate with reported skin health.

##### 3.4 Sample disposal

Nappies will be disposed on the same day in the Westwood Nursery nappy bin. Faecal material will be disposed of with other laboratory infectious waste and autoclaved.

##### 3.5 Nursery follow-up

At study end, a poster for display in the nursery and information sheets of study results will be made available to parents and staff. Papers published on the study will be shared with parents and nursery staff.

##### 3.6 Data security

Screening logs will be kept at Westwood Nursery. Paper data collection forms (CRFs) will be stored in a locked cabinet at the research office. Source data will be stored in accordance with the NHS code of confidentiality.

Research staff will ensure that the participants' anonymity is maintained through protective and secure handling and storage of participant information at the study centre. The participants will be identified only by a study ID number on the CRF and database. All documents will be stored securely and only accessible by study staff and authorised personnel. Data will be collected and retained in accordance with the Data Protection Act 2018.

Data identified by the participant's unique study number will be entered directly into the database by the research staff.

Study documents (paper and electronic) will be retained in a secure location during, and after, the study has finished. All essential documents, including participant records and other source documents will be retained for a period of 5 years following the end of the study.

### 4. Participant entry

#### Inclusion Criteria

•Any age baby aged 6-18 months at study start with parents willing to consent.

•Wears nappies 24h / day.

•Wears disposable nappies.

**Exclusion criteria**

•Any baby who, at Nursery, wears reusable nappies.

•Any baby whose parents do not consent.

• Any baby whose parent's English is not sufficient to properly understand the study.

#### 4.3 Data collection and analysis

*Who is responsible for data collection?* Data will be collected by nursery staff with the Baby room lead, Mrs Amanda Vick having overall responsibility. The form is shown in figure 3 and will be a paper form. Data will be securely stored in a locked draw in the office of the CI. Information will be transferred to an Excel spread sheet which will be securely stored on the University of Bath server. Source data will be retained for 5 years in the office of the CI, Professor Toby Jenkins.

#### 4.4 Sample size The study will aim to recruit 12 babies. It is a pilot study, and not statistically powered.

**4.5 Results dissemination** Results will be publicly disseminated by publication in the medical-scientific literature and presentation at the annual European Society for Paediatric Infectious Disease conference.

### 5. Reporting

#### 5.1 Safety reporting, Adverse Events (AE) or Adverse Reactions (AR) (AE/AR)

The stud collects soiled nappies and information on baby skin health – the participants receiving standard care and no extra intervention, drug, or procedure. There ae no foreseeable safety concerns.

#### 5.2 (Serious) Breaches

The Investigator and the research team have a responsibility to ensure that the research is conducted in accordance with the Protocol and Good Clinical Practice. Where there is a breach, this must be assessed by the Investigator and reported to the Sponsor within 24 hours of becoming aware of the event (unless it is the Sponsor that has identified the breach). Any non–serious breaches will be filed in the Investigator Site File and Trial Master File.

For serious breaches, the Ethics committee **must be notified within 7 days of the breach being identified**. The University will liaise with the research team in order to make the required notification.

**5.3 Progress reporting and end of study reporting**

The Chief Investigator will ensure that the relevant annual reports will be produced and submitted as required. The Chief Investigator will notify the REC of the end of the study. An annual progress report (APR) will be submitted to the REC within 30 days of the anniversary date on which the favourable opinion was given, and annually until the study is declared ended. If the study is ended prematurely, the Chief Investigator will notify the REC, including the reasons for the premature termination.

Within one year after the end of the study, the Chief Investigator will submit a final report with the results, including any publications/abstracts, to the REC.

##### 5.4 Final report and dissemination to parents and Nursery staff

A lay report written for parents will be disseminated via Westwood Nursery by 6 months form project end.

##### 5.5 Peer reviewed scientific publication

The team plan two peer reviewed publications: 1. A publication of the study design (by June 2024) and 2. A scientific paper on the study findings by 12 months from project end (December 2025).

### 6. Regulatory reviews

#### 6.1 Authorisations and Research Governance Statement

The study will be performed subject to favourable opinion/ authorisation/permission from all necessary regulatory and other bodies. This includes, but is not limited to, REC, HRA, sponsor and participating site.

Before the start of the study, a favourable opinion will be sought from the NHS REC.
Substantial amendments that require review by NHS REC will not be implemented until that review is in place and other mechanisms are in place to implement at site.
All correspondence with the REC will be retained.

Before any site can enrol patients into the study, the Chief Investigator/Principal Investigator will ensure that appropriate approvals from participating organisations are in place.

For any amendment to the study, the Chief Investigator, in agreement with the sponsor will submit information to the appropriate body in order for them to issue approval for the amendment. The Chief Investigator will work with sites so they can put the necessary arrangements in place to implement the amendment to confirm their support for the study as amended.

The CI has a GCP certificate (August 2023).

#### 6.2 Consent

Consent to this study is voluntary; participants will be offered appropriate written information and the opportunity to ask any study related questions before signing written consent.

##### 6.3 Confidentiality

All data and samples will be anonymised and labeled with the allocated study number along with storage details. Once testing is completed, the samples will be destroyed.

##### 6.4 Indemnity

This is a University of Bath-sponsored research study. The University of Bath has arranged Public Liability insurance to cover the legal liability of the University as Research Sponsor in the eventuality of harm to a research participant arising from management of the research by the University.

The University of Bath holds Professional Indemnity insurance to cover the legal liability of the University as Research Sponsor and/or as the employer of staff engaged in the research, for harm to participants arising from the design of the research, where the research protocol was designed by the University.

The University of Bath's Public Liability and Professional Indemnity insurance policies provide an indemnity to our employees for their potential liability for harm to participants during the conduct of the research.

##### 6.5 Sponsor The University of Bath will sponsor the study.

##### 6.6 Funding The Annette Trust is providing funding for the PhD student who will process the data in the study. We have applied for additional funding for consumables from the Rosetree Trust, but study is not contingent on obtaining this.

##### 6.7 Monitoring and Audit

The study will be monitored by the PI, Mrs Amanda Vick at Westwood nursery and the CI Professor Toby Jenkins. The PI will monitor recruitment and retention of children as well as compliance with data protection and child health monitoring. The CI will monitor the overall project and all non-nursery related work, including the laboratory studies and data analysis / dissemination. All trial-related documents will be made available, on request, for monitoring and audit by the sponsor. The monitoring plan will be developed and agreed by the sponsor.

##### 6.8 Study Management

The day-to-day management of the study will be coordinated by Dr June Mercer-Chalmers in a team with the CI, LI and Baby room lead.

#### 7. Publication policy: Peer-reviewed scientific journals, internal reports and conference presentations.

Study results will be published in the scientific literature, study design and results (as discussed in 4.5 and 5.5); a lay report will be written for parents and nursery staff. It is planned to orally disseminate results of the project in autumn 2025 at the bi-annual IMechE *Innovating for Continence* as well as the annual European Society for Pediatric Infectious Disease conference. Moreover, study results will be published in the PhD thesis of the research student, Krystal le Doare, likely in 2026. Study data will be made publicly available. Data will be owned by the University of Bath.
